# Depression Symptoms During the COVID-19 Pandemic among Well-educated, Employed Adults with Low Infection Risks

**DOI:** 10.1101/2021.01.26.21250558

**Authors:** Duncan Thomas, Tyson Brown, Donald H. Taylor, Ralph Lawton, Victoria K. Lee, Menna Mburi, Michelle Wong, Rachel Kranton

## Abstract

Levels and distributions of depression symptoms 8-10 months after the onset of the COVID-19 pandemic are reported in a population of faculty, staff, and students at Duke University who faced minimal infection and economic disruption due to the pandemic. Almost 5,000 respondents age 18-81 years who completed the 20-item Center for Epidemiological Studies-Depression (CES-D) battery reported high rates of depression symptoms with more than 40% reporting levels that indicate risk of moderate depression and 25% indicating risk of severe depression. There is a very steep age gradient with the highest levels reported by the youngest respondents of whom over 40% are at risk of severe depression. Symptoms are worse among those who report the demands of work often interfere with family responsibilities but these pressures neither explain the high reported rates nor the steep age gradient. Severe depression risks are highest among students. High levels of depression symptoms during the pandemic appear to be persistent and not confined to those at greatest risk of infection or economic insecurity.

## 1. INTRODUCTION

COVID-19 has fundamentally altered how people live and work. Beyond potential infection, since March 2020 people have been subject to stay-at-home orders as well as school and workplace closures intended to contain virus transmission. Research has documented high rates of depression symptoms at the onset of the pandemic that are associated with infection risk and economic insecurity.^1^ This study documents that ten months into the pandemic, an adult population of well-educated, employed workers with low infection rates report very high rates of depression symptoms, and these rates are highest among the youngest (18-24y).

Elevated rates of poor psychological health have been observed during the pandemic across the globe with the highest rates reported among those groups at greatest risk pre-pandemic. These include females, less-educated, lower income and urban individuals.^2,3,4,5^ Comparing pre-pandemic to the beginning of the pandemic in U.S. samples, higher rates have been documented, particularly among young adults, individuals with less education, and those who have experienced job loss or reduced income.^1^ In contrast, high rates of anxiety and stress have been documented for adults age ≥65y for whom infection and mortality rates are higher.^7^

The psychological health of the U.S. population ten months into the pandemic, after an extended period of living with limited social interaction and workplace and school closures, is not well understood. Even less is known about the psychological health of those who have been less affected by the direct impacts of COVID-19. This study is designed to address these gaps.

We report depression symptoms measured in the last quarter of 2020 in a population of adults who faced very low risks of infection, had access to health insurance, and bore little individual job loss. By focusing on this population, the study establishes that high rates of depression symptoms during the pandemic is a widespread population health concern and not limited to those with current economic insecurity or high infection risk. Moreover, because direct COVID-19 effects can be ruled out, we investigate alternative mechanisms that potentially undergird variation in depression symptoms including work characteristics, work-family interference, and interpersonal problems.

## 2. METHODS

The study was approved by the Duke University Institutional Review Board. Participants were invited by email. All participants provided electronic informed consent prior to answering any questions. The study followed the American Association for Public Opinion Research ([AAPOR](https://www.aapor.org/Publications-Media/AAPOR-Journals/Standard-Definitions.aspx)) reporting guideline.

### 2.1 Population

The target population includes all faculty, staff, and students who, in August 2020, were employed by Duke University to conduct or support research and/or teaching. The target population does not include students not employed by the university at that time, general service providers (such as security, custodial or grounds staff), or any employee who works solely for Duke University Health System.

Drawing on the university master email list, individual-specific email invitations were sent to 19,273 target subjects in October 2020, followed by three reminders over four weeks. Data were collected up to the end of December 2020. Campaigns to encourage participation included a web site, posters, social media, and follow-up emails to specific sub-groups. Of the targets, 6,938 (36%) answered at least part of the survey after completing the informed consent which explained the study goals, that participation is voluntary, the survey length (15 minutes) and data management protocols. 578 (3%) chose to opt out of participation. The analysis sample comprises 4,992 people (72% of respondents) who reported depression symptoms which were assessed on the last of 26 screens of the survey. On average, 8 questions were displayed per screen and respondents were able to go back to revise responses. Respondents were entered into a random drawing for a token gift. The instrument, which was extensively pre-tested, was deployed in Qualtrics. Cookies assured each respondent completed the survey once and data were stored on a password-protected network accessed through a VPN.

### 2.2 Key Definitions

#### Depression Risk

Depression symptoms were measured with the 20-item Center for Epidemiological Studies-Depression (CES-D) battery,^8^ asking how often the respondent experienced each symptom during the prior week on a 4-point scale: 0 (rarely or none of the time); 1 (some of the time); 2 (moderately or much of time time) and 3 (most or almost all the time). The total score (out of 60) is the primary outcome with higher scores indicating worse symptoms. The total score is complemented with two widely-used cut-offs for elevated risk of moderate (≥16) and severe (≥23) depression.^9,10^ Secondary outcomes are four sub-scales that indicate depressed affect, low positive affect, somatic activity, and interpersonal problems.^8,11^ The CES-D battery and items in each sub-scale are displayed in the Appendix.

#### Work-Life Balance

Respondents were asked how often the demands of their work interfered with their family life over the previous 4 weeks (often, sometimes, rarely, never). A binary indicator for “often” is constructed and interpreted as indicating high levels of difficulty balancing work and family life.

#### Demographic Characteristics

Age is measured in years. Gender was not restricted to either male or female but since less than 0.5% of the sample reported anything else, we use a binary indicator for males who are contrasted with females plus others. Derived from respondents’ self-reports, the race indicators are White, Black/African American, and a third category which includes Asian and those who filled in another self-description or selected more than one designation. Hispanicity is measured by a binary indicator. Three education indicators are constructed: not completed college, completed college or more, and completed a doctorate.

#### Types of University Employees

Three mutually exclusive groups of employees are identified: faculty, students, and staff. Those whose primary appointments are in the medical school are distinguished from all other university employees.

### 2.3 Statistical Analysis

After describing the sample, bivariable relationships between the CES-D score and age are presented separately for males and females using locally weighted smoothed scatterplot regressions (LOWESS)^12^ to illustrate the shape of the relationships with minimal parametric restrictions. LOWESS estimates are also displayed for moderate and severe depression risks. Magnitudes of differences are reported for 10y age groups by gender with 95% confidence intervals robust to arbitrary heteroscedasticity. Multivariable ordinary least squares (OLS) regressions document associations between the outcomes (CES-D score and 4 sub-scales) and covariates (demographics and position in the university). Parallel models are reported for binary indicators for risks of moderate and severe depression. Multivariable models with the same covariates are reported for the binary indicator that work often interfered with family. The CES-D models are extended by including that indicator as an additional covariate in order to understand the association between work-family balance and depression. The binary outcomes are estimated as linear probability models with coefficients (multiplied by 100) indicating percentage differences. Item non-response rates are very low (<0.1%) and included in the reference category (for indicator covariates) or assigned the sample average (for age). All models report heteroskedasticity-consistent 95% confidence intervals. All data analysis is conducted using STATA 16.

## 3. RESULTS

Table 1 summarizes the sample characteristics (panel A) and outcomes (panel B). The average age of respondents is 41.4 years with a range of 18 to 81 years. One third of the sample is male. Three-quarters of the sample report themselves as white, about 10% as Black/African American, with 15% in the third category; 10% of the sample report being Hispanic. Relative to the general population, the sample is well educated: over 25% have completed a doctorate, over 85% have completed at least a 4-year college degree. Of the 15% of respondents without a college degree, one-third are undergraduates studying for that degree while working for Duke University.

**Table 1.**
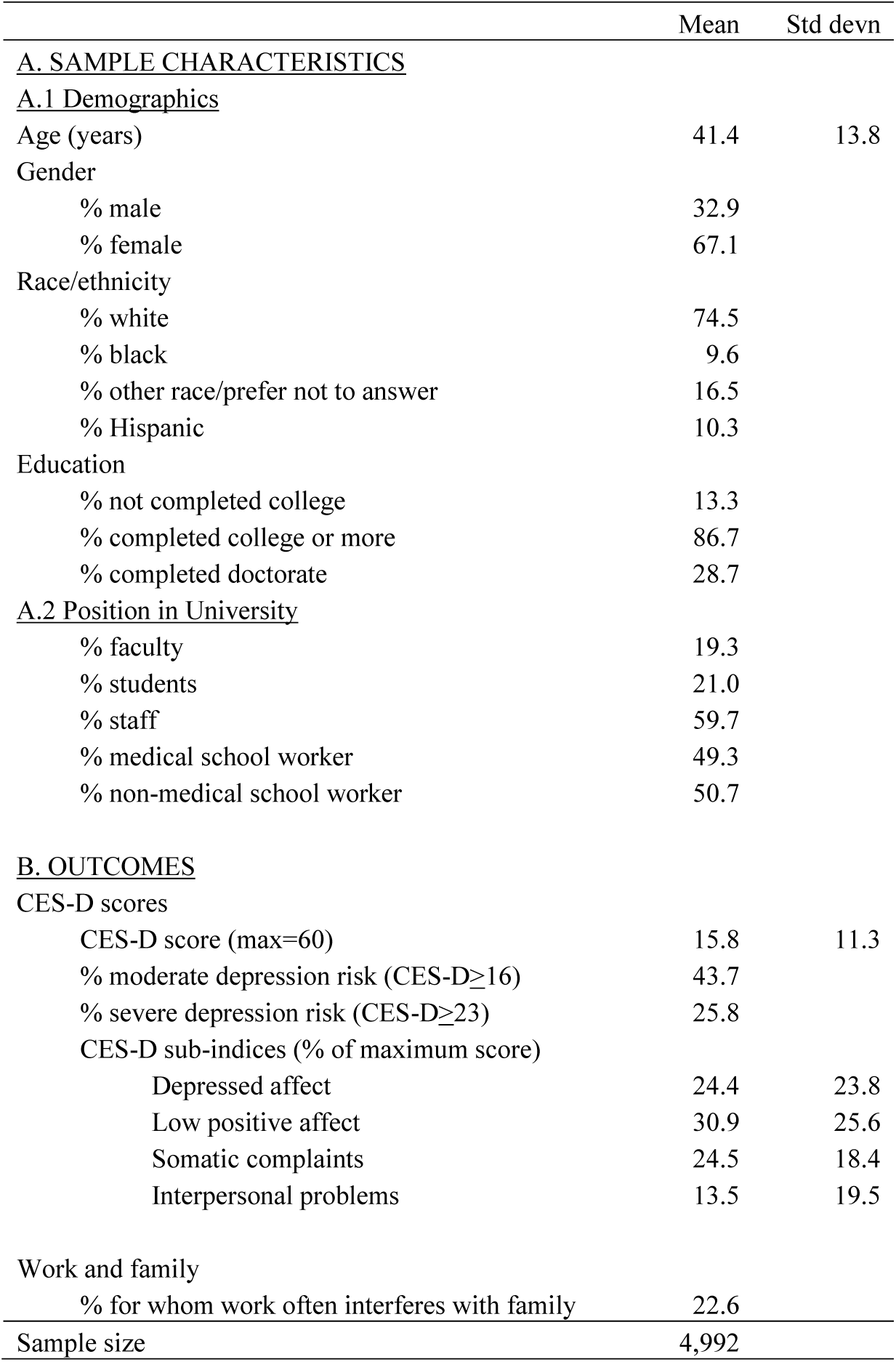
Sample characteristics and outcomes

As shown in the lower part of panel A, almost a fifth of the sample are faculty, another fifth are student-workers and the rest are staff. The medical school is the primary appointment for about half the respondents.

Outcomes are displayed in panel B of the table. On average, the CES-D score is 15.8 with over 40% of respondents reporting a score indicating risk of moderate depression, and over 25% reporting a score indicating risk of severe depression. The sub-scales establish that these high rates of symptoms are driven, almost equally, by depressed affect, low positive affect, and somatic complaints with interpersonal problems making a substantially smaller contribution. Almost one-quarter of the sample report work often interferes with family.

### Age gradients

Non-parametric estimates of the gender-specific relationship between CES-D score and age are displayed in the upper panel of Figure 1. The steep age gradient is essentially parallel for males and females with females having worse symptoms. The same pattern characterizes the percentage above the moderate and severe risk cut-offs, displayed in the lower panel of the figure, indicating very high risks of depression particularly among the youngest in the sample.

**Figure 1.**
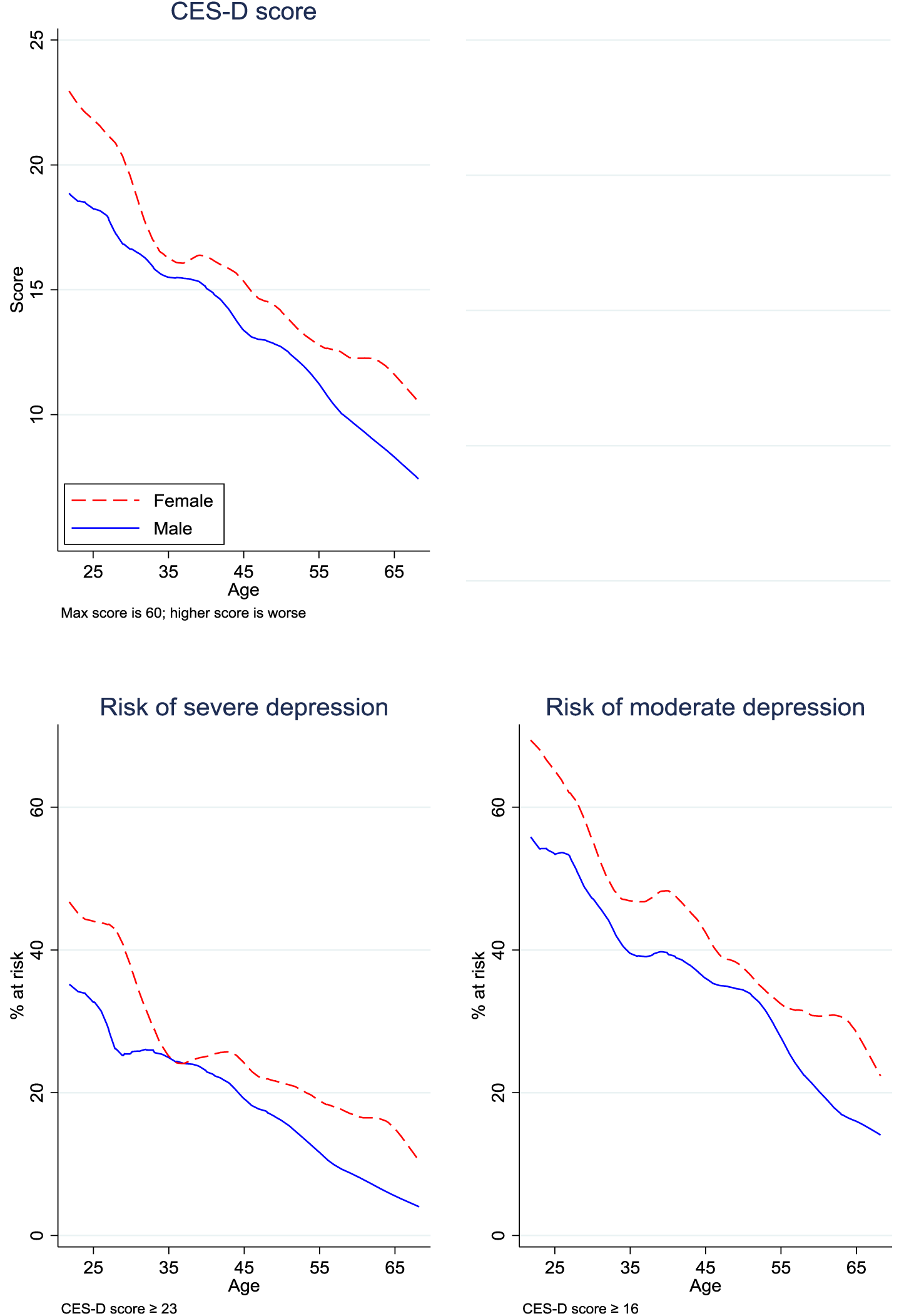
CES-D score and depression risks by age and gender

Table 2 reports the magnitudes by 10-year age groups. CES-D scores of the youngest group are over 10 percentage points higher than the oldest group. All differences between the age groups are statistically significant except for the two oldest groups. For each age group, females report significantly higher rates of symptoms than males. Over two-thirds of females and over half males age <25y report symptoms that place them at risk of moderate depression and almost half of the females and over one-third of the males are at risk of severe depression. Risks decline approximately linearly with age but remain high even among the oldest age group (≥65y), over 20% of whom report symptoms above the moderate risk cut-off.

**Table 2.**
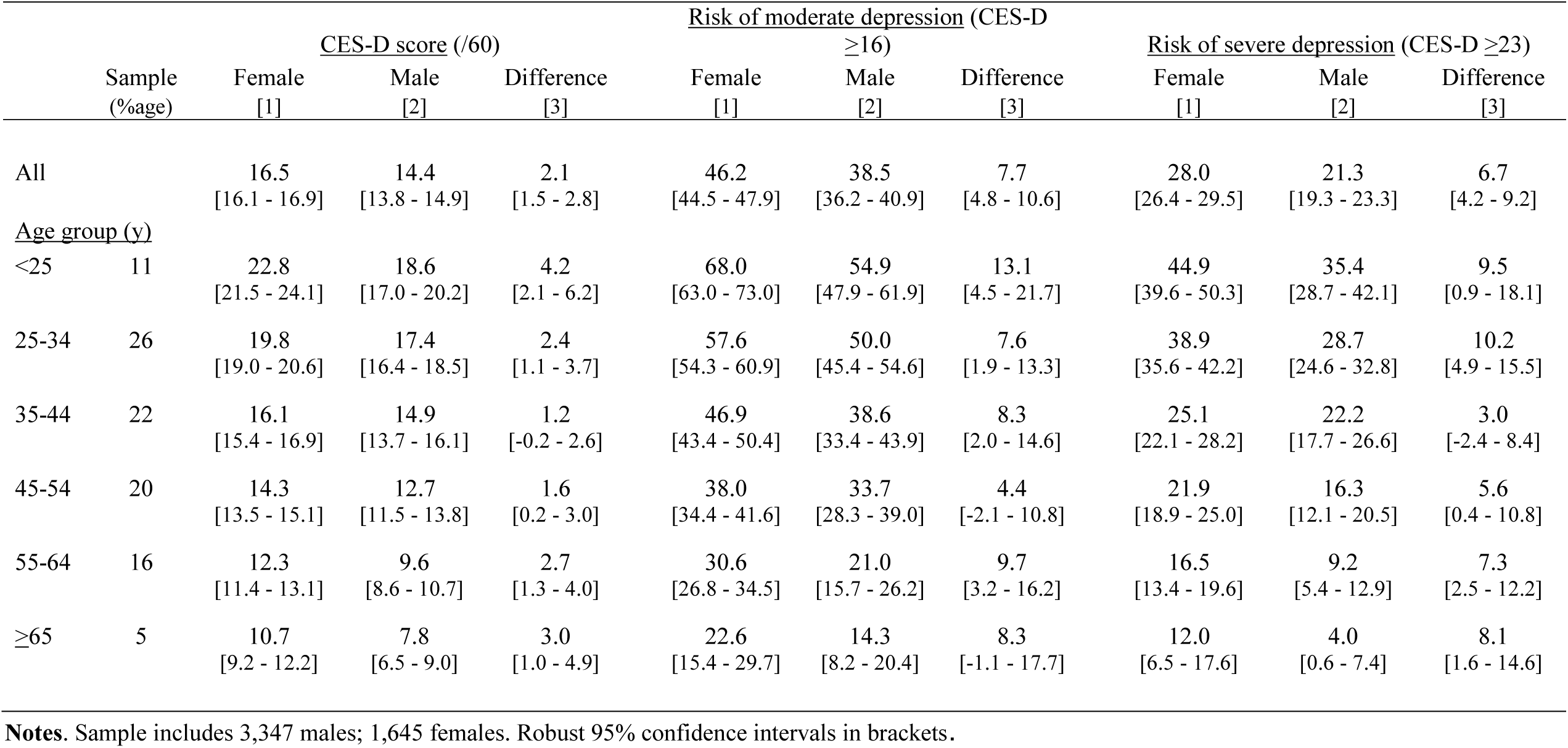
CES-D score, depression risks by age and gender

### Demographic characteristics

Multivariable regression results are reported in Table 3 with the estimated coefficients presented above 95% robust confidence intervals. The age gradient is captured with a linear term that has been demeaned so the intercept measures the level of the dependent variable for a respondent of average age. (Models that specify age as a linear spline with a knot at mean age do not reject linearity.) As indicated in Figure 1, the age gradient is steeper for risk of moderate depression relative to risk of severe risk depression. Relative to the overall score, the age gradient is significantly steeper for depressed affect, low positive affect, and somatic complaints.

**Table 3.** CES-D score, risks of moderate and severe depression and CES-D sub-indices

| Covariates | <u>CES-D score</u> | <u>Depression risks (by type)</u> |  | <u>CES-D sub-indices (% of maximum)</u> |  |  |  |
| --- | --- | --- | --- | --- | --- | --- | --- |
| | (Max=60) | Moderate<br>(CES-D ≥ 16) | Severe<br>(CES-D ≥ 23) | Depressed<br>affect | Low positive<br>affect | Somatic<br>complaints | Interpersonal<br>problems |
|  | [1] | [2] | [3] | [4] | [5] | [6] | [7] |
| <u>1. Demographics</u> |  |  |  |  |  |  |  |
| Age (years) | -0.21<br>[-0.24 - -0.19] | -0.80<br>[-0.91 - -0.68] | -0.53<br>[-0.63 - -0.43] | -0.39<br>[-0.45 - -0.34] | -0.35<br>[-0.41 - -0.29] | -0.33<br>[-0.37 - -0.29] | -0.22<br>[-0.27 - -0.18] |
| (1) if male | -2.53<br>[-3.18 - -1.89] | -9.27<br>[-12.17 - -6.36] | -8.25<br>[-10.81 - -5.69] | -6.63<br>[-7.97 - -5.29] | -2.47<br>[-3.99 - -0.95] | -4.48<br>[-5.54 - -3.42] | -1.03<br>[-2.21 - 0.16] |
| (1) if black | -0.43<br>[-1.48 - 0.63] | -1.38<br>[-5.85 - 3.08] | -1.04<br>[-5.09 - 3.02] | -0.70<br>[-2.98 - 1.58] | -1.44<br>[-3.81 - 0.94] | -0.93<br>[-2.63 - 0.76] | 0.66<br>[-1.31 - 2.63] |
| (1) if other race | -2.83<br>[-3.67 - -2.00] | -11.22<br>[-14.91 - -7.53] | -7.67<br>[-10.98 - -4.37] | -5.92<br>[-7.70 - -4.14] | -0.85<br>[-2.87 - 1.16] | -6.09<br>[-7.46 - -4.72] | -1.67<br>[-3.24 - -0.11] |
| (1) if Hispanic | 1.12<br>[0.08 - 2.15] | 0.52<br>[-3.94 - 4.98] | 4.95<br>[0.73 - 9.16] | 2.53<br>[0.28 - 4.79] | 3.47<br>[1.12 - 5.81] | 0.57<br>[-1.11 - 2.25] | 0.54<br>[-1.29 - 2.37] |
| (1) if completed college<br>or more | 0.87<br>[-0.09 - 1.83] | 3.99<br>[-0.00 - 7.98] | 2.16<br>[-1.50 - 5.83] | 0.77<br>[-1.29 - 2.83] | 2.49<br>[0.32 - 4.67] | 1.68<br>[0.12 - 3.25] | -1.22<br>[-2.93 - 0.48] |
| (1) if completed doctorate | 0.17<br>[-0.71 - 1.04] | 0.28<br>[-3.77 - 4.32] | 0.09<br>[-3.42 - 3.59] | -0.18<br>[-2.05 - 1.69] | 0.87<br>[-1.21 - 2.95] | 0.29<br>[-1.16 - 1.75] | -0.84<br>[-2.41 - 0.73] |
| <u>2. Position in university</u> |  |  |  |  |  |  |  |
| (1) if faculty | 0.04<br>[-0.91 - 1.00] | 1.97<br>[-2.51 - 6.46] | 0.53<br>[-3.27 - 4.32] | -0.30<br>[-2.32 - 1.71] | -0.87<br>[-3.19 - 1.45] | 0.79<br>[-0.79 - 2.37] | 1.01<br>[-0.73 - 2.75] |
| (1) if student | 2.65<br>[1.58 - 3.71] | 10.20<br>[5.64 - 14.75] | 10.78<br>[6.41 - 15.14] | 5.43<br>[3.14 - 7.71] | 2.71<br>[0.32 - 5.11] | 4.55<br>[2.82 - 6.27] | -0.23<br>[-2.22 - 1.75] |
| (1) if medical school worker | -2.08<br>[-2.70 - -1.45] | -7.01<br>[-9.84 - -4.18] | -6.12<br>[-8.61 - -3.63] | -3.60<br>[-4.92 - -2.27] | -5.07<br>[-6.53 - -3.60] | -2.42<br>[-3.44 - -1.40] | -0.31<br>[-1.44 - 0.83] |
| Intercept | 16.67<br>[15.63 - 17.71] | 46.06<br>[41.63 - 50.48] | 28.11<br>[24.11 - 32.10] | 27.42<br>[25.19 - 29.66] | 31.29<br>[28.90 - 33.68] | 25.61<br>[23.91 - 27.31] | 15.27<br>[13.42 - 17.13] |

**Notes.** Robust confidence intervals in brackets. Coefficients in linear probability models for depression risk models (cols 2 and 3) multiplied by 100.

Paralleling results in Figure 1 and Table 2, overall males report significantly lower symptoms than females and are less likely to score above either the moderate or severe risk cut-offs. The same is true for those who self-report as other than white or Black/African American. Hispanics report more symptoms overall and are at higher risk of severe depression. Education is not associated with levels of symptoms overall or risks of depression although those who have completed college or more score significantly worse on low positive affect and somatic complaints. Completing more than a college degree is not significantly associated with any difference in sub-scale scores.

### Type of university employment

The association between depression symptoms and type of university position, controlling for demographic characteristics, is reported in the lower half of Table 3. Relative to staff, the reference group, neither levels of symptoms nor depression risks are different among faculty. However, students report significantly more symptoms and have 10 percentage points higher risk of moderate and severe depression, adjusting for age and other demographics. University employees whose primary appointment is in the medical school report significantly lower rates of symptoms (except interpersonal problems) and are at lower risk of moderate or severe depression.

### Work-life balance

With most respondents working from home, day care centers closed and schools operating remotely, the pandemic entails difficulties balancing demands of work and family, which could underlie the patterns of depression symptoms. The first column of Table 4 reports multivariable regression estimates of the relationship between whether a respondent reported that work often interferes with family and the same covariates in Table 3. The regression model is specified as a linear spline with a knot at mean age, 41y. Work is significantly more likely to interfere with family as respondent age increases until age 41y when it declines rapidly with age. These demands are greater among females, Hispanics, and those who have completed college or more. They are also greater among faculty and students, relative to staff.

**Table 4.** CES-D score, depression risks and work/family demands

| Covariates | Work | CES-D score | Depression risks by type |  |
| --- | --- | --- | --- | --- |
| | often interferes<br>with family<br>[1] | (Max=60)<br>[2] | Moderate<br>(CES-D ≥16)<br>[3] | Severe<br>(CES-D ≥23)<br>[4] |
| <u>1. Work and family</u> |  |  |  |  |
| (1) if work often interferes<br>with family |  | 5.44<br>[4.69 - 6.19] | 21.05<br>[17.84 - 24.25] | 15.19<br>[12.11 - 18.27] |
| <u>2. Demographics</u> |  |  |  |  |
| Age (linear) |  | -0.20<br>[-0.22 - -0.17] | -0.74<br>[-0.86 - -0.62] | -0.49<br>[-0.59 - -0.39] |
| Age spline<median | 1.10<br>[0.84 - 1.36] |  |  |  |
| Age spline ≥median | -2.10<br>[-2.47 - -1.72] | | | |
| (1) if male | -3.91<br>[-6.41 - -1.41] | -2.31<br>[-2.94 - -1.68] | -8.36<br>[-11.20 - -5.51] | -7.67<br>[-10.20 - -5.15] |
| (1) if black | 0.48<br>[-3.32 - 4.28] | -0.49<br>[-1.52 - 0.54] | -1.60<br>[-6.02 - 2.82] | -1.22<br>[-5.25 - 2.81] |
| (1) if other race | -3.34<br>[-6.60 - -0.09] | -2.65<br>[-3.47 - -1.84] | -10.50<br>[-14.14 - -6.87] | -7.24<br>[-10.51 - -3.97] |
| (1) if Hispanic | 4.78<br>[0.85 - 8.72] | 0.88<br>[-0.13 - 1.90] | -0.36<br>[-4.78 - 4.05] | 4.29<br>[0.10 - 8.47] |
| (1) if completed college<br>or more | 4.93<br>[1.94 - 7.92] | 0.41<br>[-0.53 - 1.35] | 2.21<br>[-1.72 - 6.15] | 0.96<br>[-2.67 - 4.60] |
| (1) if completed doctorate | 6.01<br>[2.35 - 9.68] | -0.18<br>[-1.03 - 0.67] | -1.07<br>[-5.04 - 2.90] | -0.86<br>[-4.34 - 2.61] |
| <u>3. Position in university</u> |  |  |  |  |
| (1) if faculty | 11.77<br>[7.50 - 16.03] | -0.64<br>[-1.57 - 0.29] | -0.72<br>[-5.10 - 3.67] | -1.35<br>[-5.09 - 2.39] |
| (1) if student | 8.90<br>[4.59 - 13.21] | 2.76<br>[1.72 - 3.81] | 10.62<br>[6.11 - 15.12] | 11.16<br>[6.83 - 15.48] |
| (1) if medical school worker | 1.35<br>[-1.07 - 3.78] | -2.16<br>[-2.77 - -1.55] | -7.29<br>[-10.07 - -4.51] | -6.36<br>[-8.82 - -3.90] |
| Intercept | 25.41<br>[21.26 - 29.56] | 16.00<br>[14.99 - 17.02] | 43.46<br>[39.10 - 47.82] | 26.21<br>[22.25 - 30.17] |

**Notes.** Robust confidence intervals in brackets. Coefficients in linear probability models for work often interferes with family (col 1) and depression risk models (cols 3 and 4) multiplied by 100.

To assess the association between competing work and family demands and depression symptoms, the models in the first three columns of Table 3 are extended by including the indicator that work often interferes with family. It is positively and significantly associated with the CES-D score, and the probability of being at risk of moderate or severe depression. For example, relative to other respondents, the 20% who report work often interferes with family are 15 percentage points more likely to report symptoms indicating risk of severe depression.

Relative to the models in Table 3, the age gradients are slightly attenuated but none of the differences is substantively important or statistically significant. There are no substantively important differences in the estimates for any of the other covariates. Work and family conflicts therefore explain neither the high levels of reported depression symptoms nor the steep age gradients.

## 4. DISCUSSION

Analyzing unique data from adults 8-10 months since the start of the pandemic, this study has four novel findings on depression symptoms.

First, high rates of depression symptoms during COVID-19 are not restricted to those at greatest direct risk of the pandemic’s physical health or economic impacts. High rates are observed in our sample of university employees. All faculty, students, and staff were at very low risk of infection. (About 1% of the sample reported having been infected.) They have access to employer-provided health insurance and faced low risks of individual job loss. Yet, the respondents exhibit very high levels of depression symptoms: overall, one-quarter of the sample report symptoms indicating risk of severe depression. These levels are far higher than observed in the general population pre-pandemic as reported by meta-analyses estimating <5% of adults were at risk of severe depression.^13^ Our findings of high rates of depression symptoms among university employees during the pandemic could pertain to similar populations, not only at other colleges and universities but more broadly to employed white-collar workers.

Second, in this population there is a steep age gradient with the youngest reporting the most depression symptoms. Among those age <25y, symptoms above the cut-off for severe and moderate depression are reported by over 1/3 and 2/3 of respondents, respectively. This finding contrasts sharply with the pre-pandemic CES-D age gradient which has been shown to be essentially flat for 18-64y adults and declines with age for those age ≥65y.^14^ After adjusting for age, student workers report worse symptoms and are at higher risk of depression than faculty or staff.

Third, interpersonal problems, as measured by the CES-D sub-scale, do not explain either the high level of depression symptoms or the age gradient. High levels of reported symptoms span all four CES-D domains, with the highest levels and steepest age gradient for depressed affect, low positive affect, and somatic activity.

Fourth, work-life balance pressures are associated with significantly elevated depression symptoms. However, they do not explain the high levels of depression symptoms reported by respondents. Moreover, the associations are largely orthogonal to respondent characteristics and do not contribute to the symptom-age gradient.

One potential explanation for the steep age gradient in this population is that increased uncertainty about the future and future employment prospects weighs more heavily on younger respondents who are early in their careers. Leveraging the specifics of the university workplace, the evidence is consistent with this explanation. Conditional on age, gender, and race, students report more symptoms placing them at substantially higher risk of moderate and severe depression. The key distinguishing feature of students is that they have a relatively short horizon with regard to time at the university.

Policies and public discourse during the pandemic have focused on minimizing infection and mitigating economic disruption. Less attention has been paid to the costs of high rates of individual depression symptoms that do not stem from these immediate factors. High rates of depression symptoms found in even this well-educated, employed study sample, particularly among young adults and student workers, suggest uncertainty and concerns about the future play a role. The evidence suggests workplace and public policies that address psychosocial problems during the pandemic are likely to substantially benefit population health.

## 5. LIMITATIONS

While an online survey is well-suited for this population who rely on electronic communication in their daily lives, particularly during the pandemic, it is the only feasible option during the pandemic. An important potential limitation of the design is that those with poorer psychosocial health may have been less like to complete the survey which would result in under-stating levels of depression symptoms in the population. Selection could also operate in the opposite direction.

To investigate potential bias from self-selection of respondents, all non-respondents were invited to participate in a post-survey follow-up with an abbreviated instrument that excluded the CES-D battery but included a general emotional health question which was also included in the full survey. Overall participation increased from 72% (who completed the full survey) to 81.4%. The percentage reporting themselves in the two worst categories of general emotional health (fair or poor) is very similar in the two samples: 22.7% and 22.1% in the full survey and extended samples, respectively. (The correlation between this indicator and the CES-D score in the full survey is 0.61.) This evidence suggests non-response bias may not be important in this study.

Another limitation of the data is that the longer-term effects of the high levels of depression symptoms during the COVID-19 pandemic are not known. Future surveys of these respondents will provide evidence on possible lasting impacts.

## 6. CONCLUSIONS

Eight to ten months into the COVID-19 pandemic, high levels of depression symptoms are reported in a sample of well-educated adults employed in a university setting with low infection risk. Over 25% of the sample report depression symptoms above the cut-off for risk of severe depression. Symptoms are especially high among younger employees and student workers.

## Data Availability

Data are available from the corresponding author.

## Appendix

CES-D battery

CES-D. For each of the following ways you might have felt, please indicate how often you felt this way during the past week

1. Rarely or none of the time (<1 day)
2. Some or a little of the time (1-2 days)
3. Occasionally or a moderate amount of time (3-4 days)
4. Most or all of the time (5-7 days)

| Item | Responses | Sub-scale |
| --- | --- | --- |
| A. I was bothered by things that usually don't bother me | 1. Rarely 2. Some 3. Occasional 4. Most of time | Somatic complaints |
| B. I did not feel like eating; my appetite was poor | 1. Rarely 2. Some 3. Occasional 4. Most of time | Somatic complaints |
| C. I felt that I could not shake off the blues even<br>with help from my family or friends | 1. Rarely 2. Some 3. Occasional 4. Most of time | Depressed affect |
| D. I felt that I was just as good as other people | 1. Rarely 2. Some 3. Occasional 4. Most of time | Low positive affect |
| E. I had trouble keeping my mind on what I was doing | 1. Rarely 2. Some 3. Occasional 4. Most of time |  |
| F. I felt depressed | 1. Rarely 2. Some 3. Occasional 4. Most of time | Depressed affect |
| G. I felt that everything I did was an effort | 1. Rarely 2. Some 3. Occasional 4. Most of time | Somatic complaints |
| H. I felt hopeful about the future | 1. Rarely 2. Some 3. Occasional 4. Most of time | Low positive affect |
| J. I thought my life had been a failure | 1. Rarely 2. Some 3. Occasional 4. Most of time |  |
| K. I felt fearful. | 1. Rarely 2. Some 3. Occasional 4. Most of time |  |
| L. I had trouble sleeping or my sleep was restless | 1. Rarely 2. Some 3. Occasional 4. Most of time | Somatic complaints |
| M. I was happy | 1. Rarely 2. Some 3. Occasional 4. Most of time | Low positive affect |
| N. I talked less than usual | 1. Rarely 2. Some 3. Occasional 4. Most of time |  |
| P. I felt lonely | 1. Rarely 2. Some 3. Occasional 4. Most of time | Depressed affect |
| Q. People were unfriendly | 1. Rarely 2. Some 3. Occasional 4. Most of time | Interpersonal problems |
| R. I enjoyed life | 1. Rarely 2. Some 3. Occasional 4. Most of time | Low positive affect |
| S. I had crying spells | 1. Rarely 2. Some 3. Occasional 4. Most of time | Depressed affect |
| T. I felt sad | 1. Rarely 2. Some 3. Occasional 4. Most of time | Depressed affect |
| U. I felt people disliked me | 1. Rarely 2. Some 3. Occasional 4. Most of time | Interpersonal problems |
| V. I could not get going | 1. Rarely 2. Some 3. Occasional 4. Most of time | Somatic complaints |

## Notes

### Competing Interest Statement

All authors are employed by Duke University. No one, other than the authors, had control over the design, implementation, analyses, interpretation of data, preparation of the manuscript, or approval of the manuscript.

### Funding Statement

Trinity College of Arts & Sciences at Duke University and the Duke Social Science Research Institute provided financial support.

### Author Declarations

Duke University Institutional Review Board

